# Aggressive Systolic Blood Pressure Management as a Risk Factor of Acute Kidney Injury in Patients with Intracerebral Hemorrhage

**DOI:** 10.64898/2026.04.28.26352001

**Authors:** Victoria E Boulware, Alexander W Bae, Dillon J Dzikowicz, Ann M Leonhardt-Caprio, Daryl C McHugh, Brandon Qualls

## Abstract

**Background:** Intensive systolic blood pressure (SBP) reduction is routinely employed to limit hematoma expansion in spontaneous intracerebral hemorrhage (ICH). However, the renal consequences of sustained aggressive SBP lowering in real-world clinical practice remain incompletely characterized.

**Methods:** We conducted a retrospective cohort study of adults admitted to the intensive care unit with spontaneous ICH between 2011 and 2023. Hourly SBP measurements over the first 7 days were standardized and clustered using *k*-Shape time-series clustering to identify distinct shape-based SBP trajectories. Acute kidney injury (AKI) was defined using Kidney Disease: Improving Global Outcomes (KDIGO) criteria. Multivariable logistic regression assessed associations between SBP trajectory cluster and AKI, adjusting for demographics, baseline illness severity, renal function, and nephrotoxic medication exposure.

**Results:** Among 233 patients (mean age 61.2±14.1 years), two distinct SBP trajectories were identified: Cluster 1 (rebound SBP trajectory), a progressive upward SBP trajectory with gradual rebound, and Cluster 2 (rapid-drop SBP trajectory), a lower SBP trajectory characterized by rapid early reduction and sustained levels below 140 mm Hg. Overall, 70.4% developed AKI of any stage. Patients of Cluster 1 (rebound SBP trajectory) had significantly higher odds of AKI compared to those of Cluster 2 (rapid-drop SBP trajectory) (adjusted OR 1.97; 95% CI, 1.03–3.78). Higher maximum nicardipine dose was independently associated with AKI (OR 1.14 per mg/h; 95% CI, 1.03–1.26). SBP trajectory cluster was not significantly associated with hematoma expansion (defined as a binary outcome based on physician-documented expansion vs. no expansion), neurological outcomes, or 1-year mortality.

**Conclusions:** In ICH patients, rapid early decline in SBP followed by relative stabilization at lower levels (<140 mm Hg) is associated with increased risk of AKI without clear neurological benefit. These findings highlight the importance of balancing cerebral hemorrhage control with renal perfusion and support cautious implementation of intensive BP targets in clinical practice.

## Introduction

Spontaneous intracerebral hemorrhage (ICH) accounts for approximately 15–30% of all strokes and remains associated with high morbidity and mortality despite advances in critical care management^1^. A recent study on 14 years of data (2004–2018) from the Nationwide Inpatient Sample revealed an overall annual ICH incidence rate of 23.15 cases per 100,000 population^2^. During this period, the total number of ICH cases in the United States increased by 11%, accompanied by a 16% rise in hypertension prevalence and a 130% increase in anticoagulant use^2^.

Elevated systolic blood pressure (SBP) after ICH is strongly associated with hematoma expansion, neurological deterioration, and poor outcomes^3^. Multiple studies have been conducted to create guidelines for the SBP target range, attempting to weigh the neuroprotective benefits against the risk of ischemic damage to other organs^4,5,6,7,8^. Meta analysis of ATACH-2 (Antihypertensive Treatment of Acute Cerebral Hemorrhage-2) trial cohort, which includes 1,000 ICH patients randomly divided into intensive and standard treatment groups based on targeted SBP (110–139 mmHg and 140–179 mmHg, respectively), demonstrated that lower SBP targets were associated with a reduced incidence of hematoma expansion. However, aggressive SBP reduction (targeted SBP 110–139 mmH) was linked to an increased risk of renal and cardiopulmonary ischemia, as well as higher morbidity. This was particularly true for patients whose SBP reduction was greater than 90 mmHg or intensive reduction in patients who had SBP at admission higher than 210 mmHg^7^. The INTERACT-2 (Intensive Blood Pressure Reduction in Acute Cerebral Hemorrhage) trial evaluated whether intensive lowering of SBP to <140 mmHg within one hour would improve outcomes in patients with acute spontaneous ICH compared with guideline-recommended management (<180 mmHg). In this multicenter randomized trial of 2,839 patients, intensive SBP lowering did not significantly reduce the composite outcome of death or major disability at 90 days^8^.

Although the 2022 Guideline for the Management of Patients With Spontaneous Intracerebral Hemorrhage: A Guideline From the American Heart Association/American Stroke Association is based on clinical trials that prospectively evaluated adverse events according to predefined SBP targets, there is a lack of retrospective studies validating these findings in the real clinical world^5,9^. This study is designed to examine actual SBP trajectories during intensive care unit (ICU) admission in real-world clinical practice.

## Method

### Study Population

The study population comprised adults aged 18–85 years, of any sex or race, who were admitted to the intensive care unit between January 2011 and December 2023 with a primary diagnosis of ICH. Eligible cases were initially identified using International Classification of Diseases, Tenth Revision (ICD-10) diagnosis codes for spontaneous ICH (I61.0-I61.9), and all identified cases were subsequently verified through registered nurse–led manual chart review to confirm the diagnosis. ICD-10 codes have a high positive predictive value (89-96%) when verified against patient notes, indicating excellent coding accuracy for ICH^10,11^. Although national guidelines for SBP management in ICH were updated in 2022, near the end of the study inclusion period, delayed changes in clinician practice resulted in a consistent blood pressure management approach throughout the study^9^.

Patients were excluded if they had preexisting chronic kidney disease (CKD) stage III or worse, including those with an admission estimated glomerular filtration rate (eGFR) ≤29 mL/min/1.73 m², due to the high baseline risk of renal dysfunction and the potential confounding effect on outcomes related to blood pressure management^12^. Patients with a Glasgow Coma Scale (GCS) score of 3–5 on admission were excluded because the severity of neurological injury in this population is associated with poor prognosis independent of blood pressure control^13^. Individuals with traumatic intracerebral hemorrhage were excluded to maintain a homogeneous cohort of spontaneous ICH, as the pathophysiology and management strategies differ substantially^14^. Finally, patients with a hospital length of stay of less than 5 days (120 hours) were excluded to ensure adequate observation of systolic blood pressure trajectories and associated clinical outcomes during the acute and subacute phases of care.

### Data collection

All data was retrieved from the electronic medical records (Epic, Verona, WI) and manually reviewed by the study team. The following information was collected: age, gender, race, past medical history of cerebrovascular accident, seizure disorder, or renal disease, SBP, diastolic blood pressure (DBP), mean arterial pressure (MAP), GCS, National Institute of Health Stroke Scale (NIHSS), ICH score, Modified Rankin Scale (mRS), creatinine, GFR, urine output (UOP), hematoma location, volume, presence of hydrocephalus, insertion of external ventricular drain (EVD), use of selected medications (nicardipine, hydralazine, labetalol, NaCl 3%, NaCl 23%, ioxehol, mannitol), highest dose of nicardipine administered, length of stay in hospital (LOS). SBP and DBP were measured either using an automatic sphygmomanometer or via an arterial line blood pressure monitor^15,16,17,18^. Measurements from both methods were included. However, in cases where simultaneous readings were available from both devices, the sphygmomanometer measurement was prioritized as we did not have arterial waveforms to access the accuracy of the waveforms^19,20^.

Volumetric hematoma measurements (e.g., ABC/2 measurements) were not consistently available in the medical record and thus quantitative assessment of hematoma expansion could not be performed. Hematoma expansion was instead determined from narrative clinical documentation, including radiology reports and provider notes (neurologists, radiologists, or neurosurgeons), indicating whether interval imaging demonstrated an increase in hematoma size. As such, hematoma expansion was classified as a binary variable (expansion vs. no expansion) based on clinical interpretation, and precise volumetric changes were not available for analysis.

Respiratory rates within 24 hours of morbidity and ventilator respiratory rates were also collected. GCS, NIHSS, and mRS were collected from written notes and assessment documentation in the medical record. If scores were not calculated on both admission and discharge, a certified registered nurse completed the calculations based on correlated neurological assessment. Complete neurological assessment information was not present for all patients, resulting in a risk for underestimated NIHSS scores, over-representing neurological recovery at discharge. BP, UOP, and lab values were collected from time of admission to 7 days (168 hours) after admission. UOP values were verified by documented placement of indwelling urethral catheter. To standardize MAP measurements, all values were calculated based on SBP and DBP data in the following formula: MAP = DBP + 1/3(SBP-DBP)^21^.

### Outcomes

The outcomes examined in this study were neurological adverse events stratified by NIHSS increase, GCS decrease, hematoma expansion, AKI based on KDIGO standards, ICH score, in-hospital mortality and 1 year mortality. Secondary outcomes were interpreted as exploratory and not adequately powered.

### Data processing

To standardize SBP measurements to hourly intervals, a sequential selection algorithm was applied to each patient’s time series (R package lubridate). The algorithm identified and retained measurements that were 50–70 minutes apart to approximate one hour sampling while preserving the temporal continuity of the original recordings. Measurements outside this interval were excluded. When temporal gaps exceeding 70 minutes were detected, the algorithm inserted NA corresponding to the number of expected hourly intervals. This approach ensured consistent temporal spacing across patients and prevented duplication of identical timestamps, resulting in a dataset of uniformly spaced (∼1 hour) SBP recordings suitable for time-series analysis.

### Time-series SBP trajectory clustering strategy

Hourly SBP trajectories over 7 days (168hrs) were clustered using the *k*-Shape algorithm (R package dtwclust). To ensure comparability across patients, each trajectory was z-normalized by subtracting the mean and dividing by the standard deviation. Clustering was performed across a range of cluster numbers (*k* = 2–6) on equal-length, z-normalized trajectories. The algorithm employed shape-based distance (SBD) and the *k*-Shape centroid update, which preserves the intrinsic shape characteristics of time-series data. For each candidate *k*, the average silhouette width was computed based on SBD, and the optimal number of clusters (*k*) was determined with the highest silhouette value^22^. Cluster robustness was further evaluated using a stability index. All data analysis was done using R version 2025.05.0 (Posit, MA, USA). <u>AKI Diagnosis</u>

AKI was defined according to the Kidney Disease: Improving Global Outcomes (KDIGO) criteria, incorporating serum creatinine and UOP parameters consistent with the Acute Kidney Injury Network (AKIN) and RIFLE (Risk, Injury, Failure, Loss, End-stage renal disease) guidelines^23,24,25^. Stage 1 AKI was defined as an increase in serum creatinine by ≥0.3 mg/dL from baseline or 1.5–1.9 times the baseline value, or a UOP of <0.5 mL/kg/h for 6–12 hours. Stage 2 AKI was defined as an increase in serum creatinine to 2.0–2.9 times baseline, or a UOP of <0.5 mL/kg/h for ≥12 hours. Stage 3 AKI was defined as an increase in serum creatinine to ≥4.0 mg/dL or ≥3 times baseline, a UOP of <0.3 mL/kg/h for ≥24 hours, anuria for ≥12 hours, or the initiation of renal replacement therapy^26^.

### Nephrotoxic Medications

Administration of potentially nephrotoxic medications was evaluated as a covariate in the multivariable logistic regression models. The following agents were included: NaCl 3%, NaCl 23%, Iohexol (iodinated contrast medium), Mannitol, and Protamine sulfate^27^. For each patient, exposure to these agents during hospitalization was coded as a binary variable based on administration records. These variables were entered as independent predictors to adjust for potential renal effects of medication exposure in the regression analyses.

### Statistical Analysis

Baseline characteristics and in-hospital treatments were summarized overall and by SBP trajectory cluster. Continuous variables were reported as mean±SD or median (IQR) depending on distribution assessed by visual inspection and Shapiro–Wilk testing. Between-cluster comparisons used Student’s t test or Wilcoxon rank-sum tests for continuous variables and χ² or Fisher’s exact tests for categorical variables.

The association between SBP trajectory cluster and AKI was estimated using multivariable logistic regression, reporting adjusted odds ratios (aOR) with 95% confidence intervals. Covariates in the models included age, sex, race, admission NIHSS, admission SBP, admission creatinine, nicardipine exposure and maximum nicardipine dose, and binary exposure to prespecified nephrotoxic agents. Similar approaches were used for the secondary outcomes including hematoma expansion, change in NIHSS and GCS score, in-hospital mortality, and 1-year mortality.

## Results

### Basic Demographics and Descriptive Statistics

A total of 233 patients were included in this study. Average age was 61.2 (SD = 14.1) and 48.5% (n=113) were identified as male. 73.8% (n=172) were Caucasian, 15.5% (n=36) were African American, 3.4% (n=8) were Asian, and 7.3% (n=17) were unknown or other. The average SBP at admission was 171 mm Hg (SD = 40 mm Hg), and the average DBP was 93.5 mm Hg (SD = 24.1 mm Hg). 64.8% (n=151) received nicardipine infusion and the average maximum dose of nicardipine was 10.7 mg/hour (SD = 4.13 mg/hour). (Table 1.) summarizes baseline demographic characteristics of the overall cohort, Cluster 1 (rebound SBP trajectory) and Cluster 2 (rapid-drop SBP trajectory). (Table 2.) summarizes the occurrence of AKI and number of patients under active BP treatment for Cluster 1 (rebound SBP trajectory) and Cluster 2 (rapid-drop SBP trajectory).

**Table 1.** Demographic characteristics of the overall cohort, Cluster 1 (rebound SBP.

|  | **Entire patients** | **Cluster 1**<br>**(rebound SBP**<br>**trajectory)** | **Cluster 2**<br>**(rapid-drop SBP**<br>**trajectory)** |
| --- | --- | --- | --- |
| **N** | N=233 | N=111 | N=122 |
| **Mean age (±SD)** | 61.2 (±14.1) | 60.5 (±14.1) | 61.8 (±14.1) |
| **White Race** | 73.8% (n=172) | 68.5% (n=76) | 78.7% (n=96) |
| **Sex** | 48.5% (n=113) male | 47.7% (n=53) male | 49.2% (n=60) male |
| **Baseline NIHSS**<br>**Average (±SD)** | 13.5 (±8.4) | 13.4 (±8.1) | 13.7 (±8.6) |
| **Baseline GCS**<br>**Average (±SD)** | 11.2 (±3.2) | 11.1 (±3.3) | 11.4 (±3.2) |
| **Average**<br>**Admission BP** | SBP: 171 mm Hg (SD<br>= 40 mm Hg)<br>DBP: 93.5 mm Hg (SD<br>= 24.1 mm Hg)<br>MAP: 120 mm Hg (SD<br>= 27.7 mm Hg) | SBP: 161.7 mm Hg<br>(SD = 38.9 mm Hg)<br>DBP: 91.4 mm Hg (SD<br>= 22.5 mm Hg)<br>MAP: 113.9 mm Hg<br>(SD = 26.3 mm Hg) | SBP: 179.7 mm Hg<br>(SD = 39.2 mm Hg)<br>DBP: 98.8 mm Hg (SD<br>= 25.0 mm Hg)<br>MAP: 125.8 mm Hg<br>(SD = 27.9 mm Hg) |

**Table 2.** Occurrence of AKI and number of patients under active BP treatment for Cluster 1 (rebound SBP trajectory) and Cluster 2 (rapid-drop SBP trajectory). *Active BP treatment was defined as whether the patient received a nicardipine infusion.

|  | **Overall Developed AKI (Stage 1-3)** | **Overall No AKI** | **Total** |
| --- | --- | --- | --- |
| **Cluster 1 (rebound SBP trajectory)** | n=70 | n=41 | n=111<br>(n=70; 63.1% was under active BP treatment*) |
| **Cluster 2 (rapid-drop SBP trajectory)** | n=94 | n=28 | n=122<br>(n=81; 66.4% was under active BP treatment*) |
| **Total** | n=164<br>(n=112; 68.3% was under active BP treatment*) | n=69<br>(n=39; 56.5% was under active BP treatment*) |  |

34.3% (n= 80) of patients expired within 1 year of admission; 75% of which (n = 60) expired during admission. Of significant note, 61.25% (n = 48) of patients that expired were transitioned to comfort care only during admission. These patients were excluded from the survival analysis. Ordered SBP ranges (Figure 1.) were generally lower than 2022 Guideline for the Management of Patients With Spontaneous Intracerebral Hemorrhage: A Guideline From the American Heart Association/American Stroke Association^9^.

**Figure 1.**
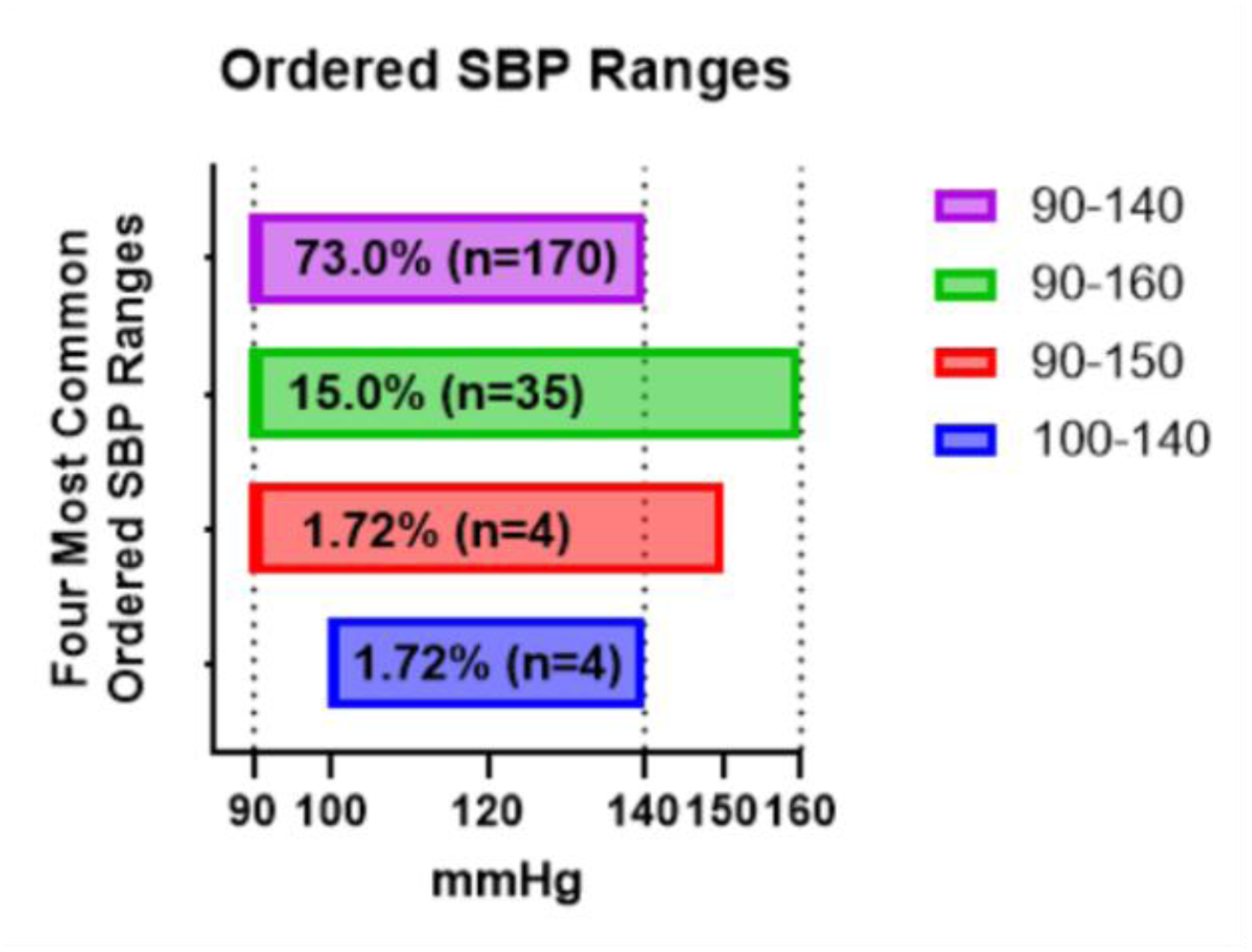
Ordered SBP ranges. 73.0% (n=170) had 90 mmHg – 140 mmHg. 15.0% (n=35) had 90 mmHg – 160 mmHg, 1.72% (n=4) had 90 mmHg – 150 mmHg and 100 mmHg – 140 mmHg.

### SBP trajectory clustering result

The *k*-Shape clustering algorithm identified two distinct clusters with a stability index of 0.85 (Figure 2.). Higher stability index (closer to 1) indicate that the identified clusters are robust to initialization and reflect stable underlying temporal patterns.

**Figure 2.**
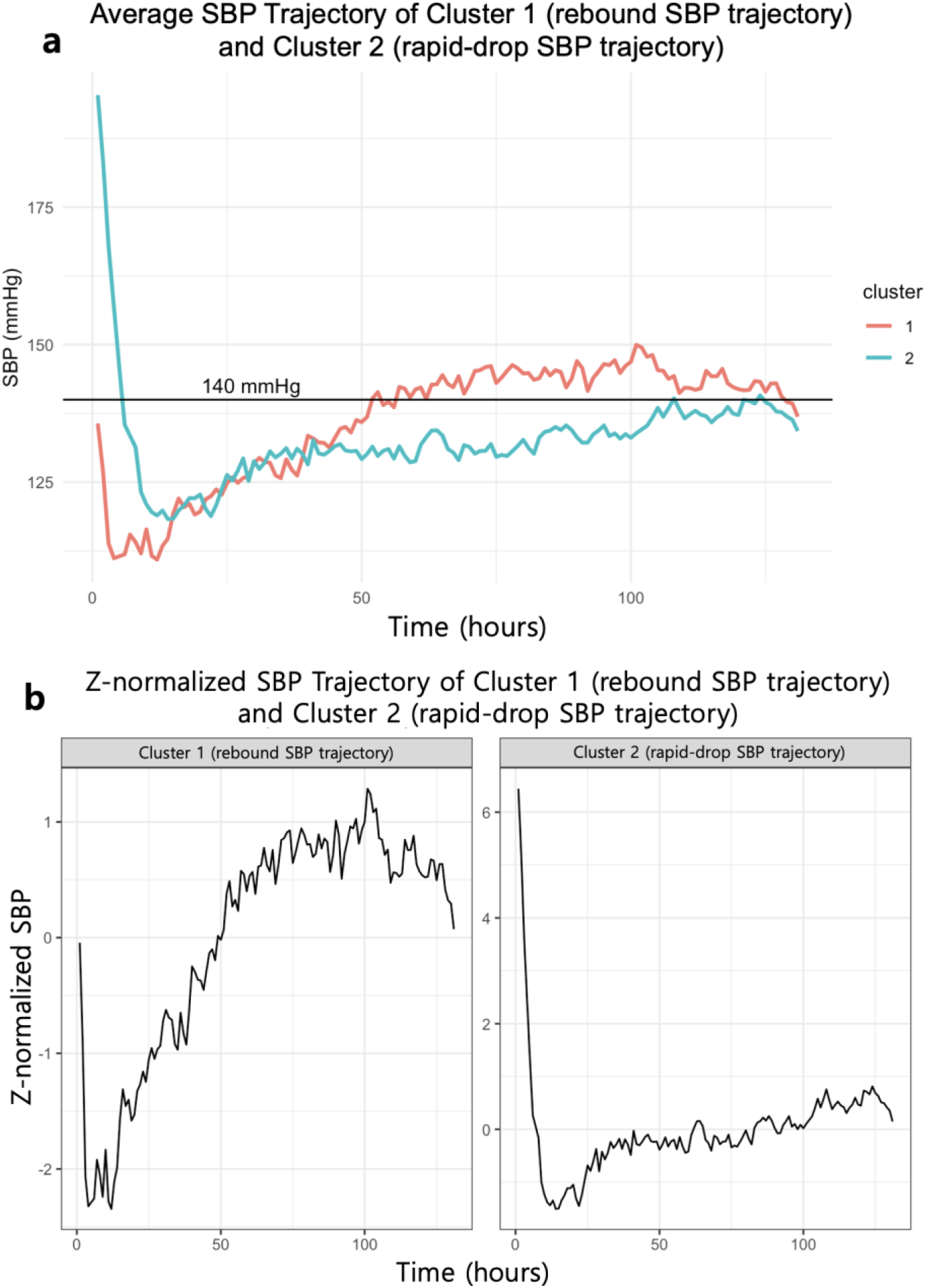
**(a)** Centroids of two SBP clusters, with Cluster 1 (red; rebound SBP trajectory) demonstrating a gradual increase above 140 mmHg and Cluster 2 (blue; rapid-drop SBP trajectory) showing an early sharp decline followed by stabilization below the 140 mm Hg threshold (black solid line). **(b)** representative z-normalized SBP trajectories for each cluster: Cluster 1 (rebound SBP trajectory) is characterized by progressive SBP elevation, whereas Cluster 2 (rapid-drop SBP trajectory) exhibits an initial sharp drop followed by relatively stable SBP.

Cluster 1 (rebound SBP trajectory) demonstrated a progressive upward trajectory, characterized by an initial decline followed by gradual SBP increase and sustained higher values over time. Cluster 2 (rapid-drop SBP trajectory) exhibited a rapid early decline in SBP followed by relative stabilization at lower levels throughout the observation period. These clusters reflect differences in temporal SBP patterns, rather than baseline SBP or treatment intensity.

Both clusters exhibited a marked decline in SBP within the first 12 hours. Cluster 1, representing the patient group with rebound SBP trajectory, showed a z-normalized SBP decrease from 0 to -2, whereas Cluster 2, representing the patient group with rapid-drop SBP trajectory, demonstrated a decrease from 6 to -1. Over time, Cluster 1 displayed a gradual increase in SBP above 140 mm Hg, while Cluster 2 showed an early, sharp decline followed by stabilization below the 140 mm Hg threshold. Cluster 1 (rebound SBP trajectory) comprised 47.6% of patients (n = 111), and Cluster 2 (rapid-drop SBP trajectory) comprised 52.3% (n = 122).

### AKI Diagnosis

Daily AKI diagnosis was made by comparing creatine level and UOP from baseline measurement. The highest stage observed across all time points was assigned as the patient’s overall AKI stage. Over the initial 7-days period following admission, 46.8% of patients had Stage 1 AKI (n = 109), 29.6% had no AKI (n = 69), 15.0% had

Stage 3 AKI (n = 35), and 8.6% had Stage 2 AKI (n = 20) (Figure 3.; Table 3.).

**Figure 3.**
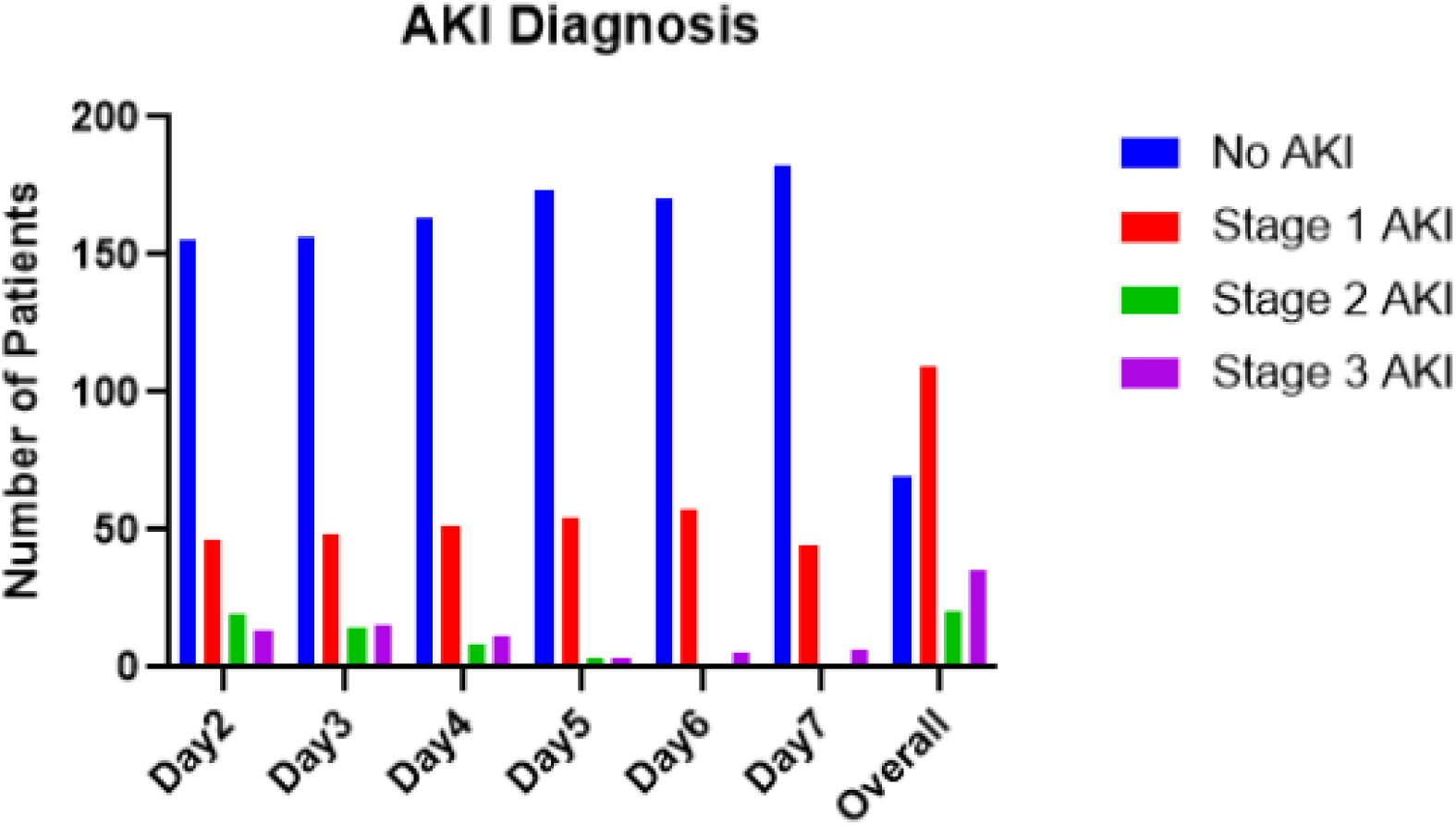
AKI diagnosis result for each day in ICU

**Table 3.** Distribution of acute kidney injury (AKI) status by hospital day. Values represent the number of patients and corresponding percentages classified as no AKI or AKI stages 1–3 on each study day (Days 2–7), demonstrating temporal changes in AKI severity over the course of hospitalization.

| **Day** | **2** | **3** | **4** | **5** | **6** | **7** | **Overall** |
| --- | --- | --- | --- | --- | --- | --- | --- |
| **No AKI** | n=157<br>(67.4%) | n=159<br>(68.2%) | n=166<br>(71.2%) | n=175<br>(75.1%) | n=173<br>(74.2%) | n=186<br>(79.8%) | n=69<br>(29.6%) |
| **Stage 1** | n=47<br>(20.2%) | n=49<br>(21.0%) | n=51<br>(21.9%) | n=56<br>(24.0%) | n=58<br>(24.9%) | n=44<br>(18.9%) | n=109<br>(46.8%) |
| **Stage 2** | n=20<br>(8.6%) | n=14<br>(6.0%) | n=9<br>(3.9%) | n=3<br>(1.3%) | n=5<br>(2.1%) | n=1<br>(0.4%) | n=20<br>(8.6%) |
| **Stage 3** | n=13<br>(5.6%) | n=15<br>(6.4%) | n=11<br>(4.7%) | n=3<br>(1.3%) | n=1<br>(0.4%) | n=6<br>(2.6%) | n=35<br>(15.0%) |

### Risk Analysis

#### AKI risk

In multivariable logistic regression analysis, SBP trajectory cluster was significantly associated with overall AKI. Patients in Cluster 2 (rapid-drop SBP trajectory) had nearly twofold higher odds of AKI compared with those in Cluster 1 (rebound SBP trajectory) (OR = 1.97; 95% CI, 1.03–3.78). Administration of 23.4% sodium chloride was associated with lower odds of AKI (OR = 0.28; 95% CI, 0.12–0.68). Other variables, including age (OR = 1.02; 95% CI, 0.999–1.05), sex (male: OR = 1.50; 95% CI, 0.74–3.04), race (Black: OR = 2.36; 95% CI, 0.87–6.37; Other: OR = 3.11; 95% CI, 0.76–12.80), baseline NIHSS score (OR = 1.03; 95% CI, 0.99–1.07), baseline SBP (OR = 1.00; 95% CI, 0.99–1.01), and baseline creatinine level (OR = 2.92; 95% CI, 0.62–13.73), were not statistically significant predictors of AKI. Similarly, the administration of nephrotoxic medications—including 3% sodium chloride (OR = 1.00; 95% CI, 0.49–2.05), iohexol (OR = 0.89; 95% CI, 0.41–1.92), mannitol (OR = 0.63; 95% CI, 0.31–1.30), and protamine sulfate (OR = 1.32; 95% CI, 0.49–3.54)—was not significantly associated with AKI risk.

However, the maximum dose of nicardipine was significantly associated with the development of AKI (OR = 1.14; 95% CI, 1.03–1.26). This indicates that each incremental increase in nicardipine dose was associated with a higher odd of AKI.

#### Hematoma expansion risk

No statistically significant association was observed between SBP trajectory cluster and hematoma expansion. Patients in the Cluster 2 (rapid-drop SBP trajectory) had higher, but not statistically significant odds of hematoma expansion compared with the Cluster 1 (rebound SBP trajectory) (OR = 2.10; 95% CI, 0.85–5.18). Other covariates, including age (OR = 1.03; 95% CI, 0.99–1.06), sex (OR = 0.95; 95% CI, 0.38–2.38), baseline SBP (OR = 1.00; 95% CI, 0.99–1.01), NIHSS score (OR = 0.96; 95% CI, 0.91–1.01), and baseline creatinine (OR = 0.49; 95% CI, 0.07–3.49), were not independently associated with hematoma expansion. Administration of 23.4% sodium chloride showed a nonsignificant trend toward higher odds of hematoma expansion (OR = 2.54; 95% CI, 0.85–7.61).

#### Discharge value of NIHSS, GCS and mRS

No statistically significant association was observed between SBP trajectory cluster and discharge value of NIHSS, GCS and mRS. Age and the admission NIHSS showed weak association with the discharge value of GCS and mRS. Age (OR = 1.08; 95% CI, 1.03–1.06) and NIHSS score (OR = 1.03; 95% CI, 1.01–1.05) were independently associated with discharge value of mRS. Age (OR = 1.08; 95% CI, 1.02–1.15) and NIHSS score (OR = 1.28; 95% CI, 1.15–1.42) were independently associated with discharge value of GCS.

#### 1 year mortality & survival analysis

During 1-year follow-up, mortality occurred in 31 of 110 patients (28.2%) in Cluster 1 (rebound SBP trajectory) and 45 of 121 patients (37.2%) in Cluster 2 (rapid-drop SBP trajectory). Kaplan–Meier survival analysis demonstrated no statistically significant difference in survival between two clusters (log-rank χ² = 1.6, p = 0.20) (Figure 4).

**Figure 4.**
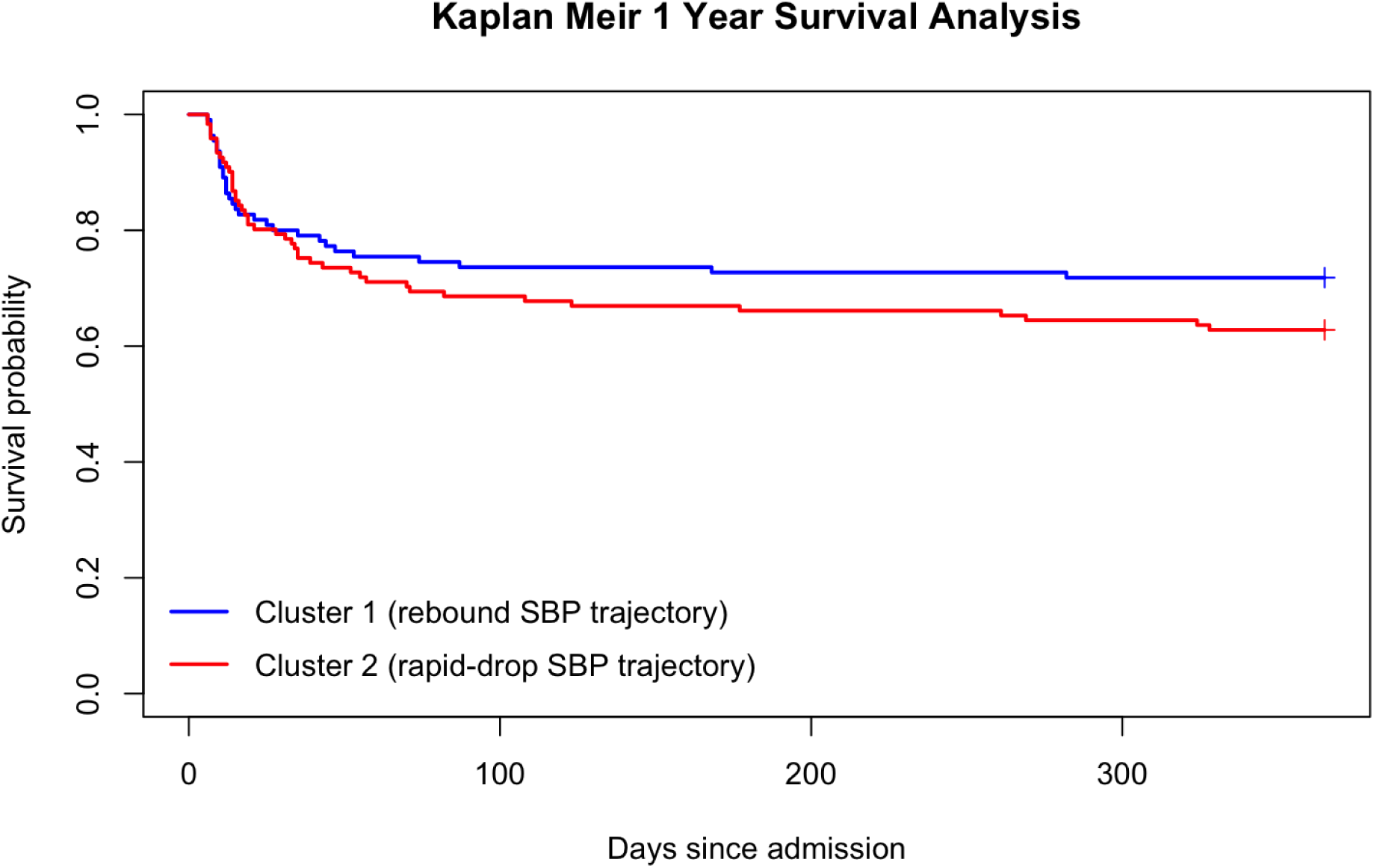
Kaplan Meier curves for 1-year all-cause mortality stratified by SBP trajectory cluster. Time zero corresponds to hospital admission. Patients alive at 1 year were censored at 365 days. Survival curves were compared using the log rank test.

Age and NIHSS scores were significant predictors of mortality. Each one-year increase in age was associated with 8.7% higher odds of 1 year mortality (OR = 1.09; 95% CI, 1.05–1.12), and each one-point increase in NIHSS score was associated with 4.3% higher odds of death (OR = 1.04; 95% CI, 1.00–1.09). Patients who received 23.4% sodium chloride also had significantly greater odds of mortality (OR = 3.61; 95% CI, 1.46–8.93), likely reflecting its use in patients with more severe disease.

Other variables—including SBP trajectory cluster (OR = 1.21; 95% CI, 0.64–2.31), sex (male: OR = 0.92; 95% CI, 0.46–1.84), baseline creatinine (OR = 0.69; 95% CI, 0.17–2.83), mannitol (OR = 1.38; 95% CI, 0.65–2.97), and race (Black: OR = 0.89; 95% CI, 0.35–2.25; Other: OR = 1.54; 95% CI, 0.46–5.20)—were not significantly associated with mortality.

In multivariable Cox proportional hazards analysis adjusting for age, sex, race, AKI stage, NIHSS, and GCS, SBP trajectory cluster was not independently associated with 1-year mortality (Low SBP vs High SBP: adjusted hazard ratio [HR] 1.14, 95% CI 0.70–1.88; p = 0.59). Increasing age was independently associated with higher mortality risk (adjusted HR per year 1.05, 95% CI 1.03–1.08; p < 0.001). Higher GCS demonstrated a borderline association with lower mortality (adjusted HR per point 0.91, 95% CI 0.83–1.00; p = 0.058).

#### In-hospital mortality

During hospitalization, death occurred in 22 of 110 patients (20.0%) in Cluster 1 (rebound SBP trajectory) and 32 of 121 patients (26.4%) in Cluster 2 (rapid-drop SBP trajectory). Kaplan–Meier analysis demonstrated no statistically significant difference in in-hospital survival between clusters (log-rank χ² = 1.4, p = 0.20) (Figure 5).

**Figure 5.**
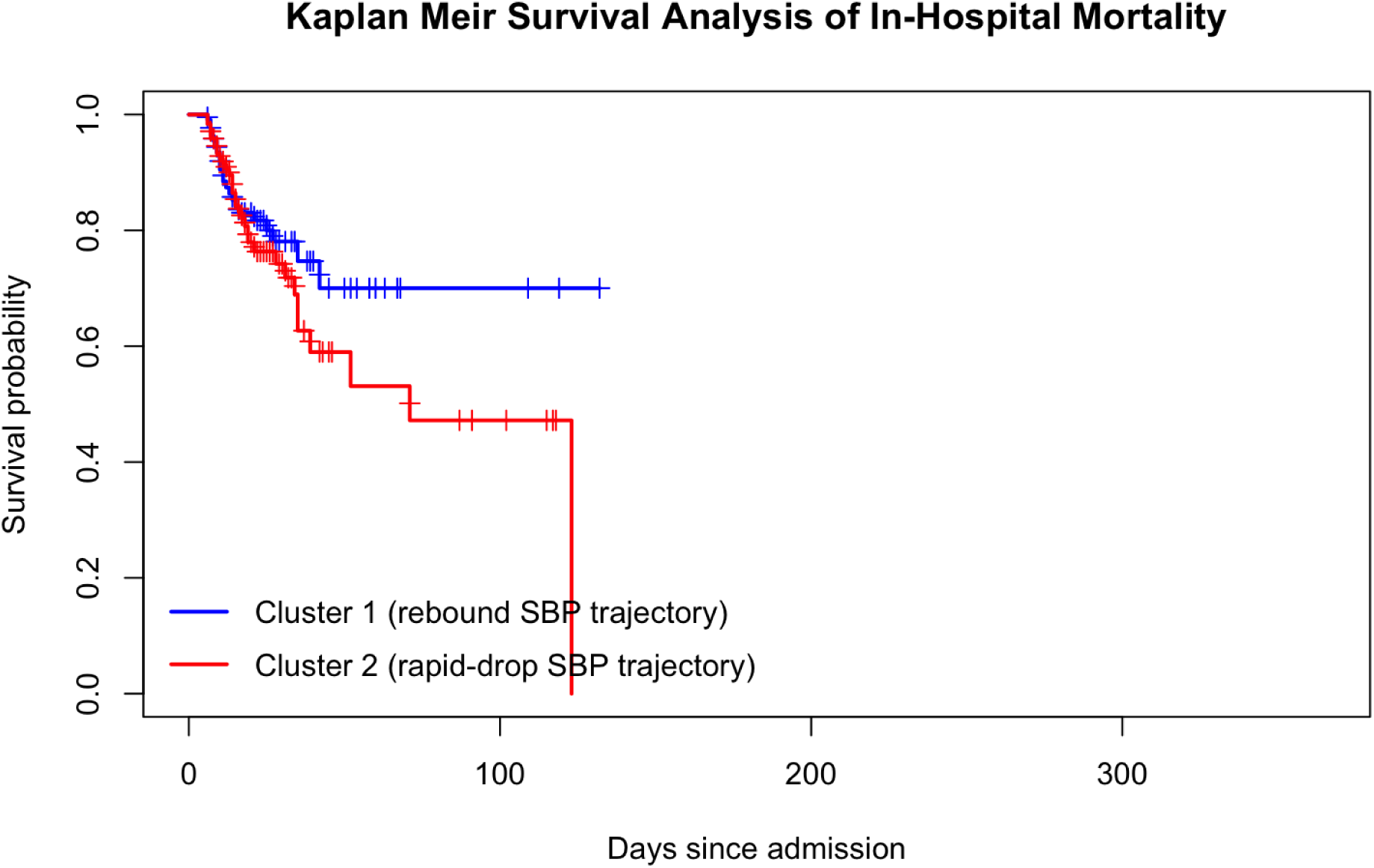
Kaplan–Meier curves for in-hospital mortality stratified by SBP trajectory cluster. Time zero corresponds to hospital admission. Patients who survived to discharge were censored at the time of discharge. Survival curves were compared using the log-rank test.

In multivariable Cox proportional hazards regression adjusting for age, sex, race, AKI stage, NIHSS, and GCS, SBP trajectory cluster was not independently associated with in-hospital mortality (Low SBP vs High SBP: adjusted HR 1.10, 95% CI 0.60–2.03; p = 0.75). Increasing age was independently associated with higher in-hospital mortality (adjusted HR per year 1.06, 95% CI 1.03–1.09; p < 0.001). Other covariates, including sex, AKI stage, race, NIHSS, and GCS, were not independently associated with in-hospital mortality.

## Discussion

In this real-world ICU cohort of adults with spontaneous ICH, we identified two reproducible SBP trajectory phenotypes over the first 7 days using shape-based time-series clustering: Cluster 1 (rebound SBP trajectory), a progressive upward SBP trajectory with gradual rebound, and Cluster 2 (rapid-drop SBP trajectory), a lower SBP trajectory characterized by rapid early reduction and sustained SBP generally <140 mm Hg. Cluster 2 (rapid-drop SBP trajectory) was independently associated with nearly twofold higher odds of AKI, while no statistically significant associations were observed between SBP trajectory and hematoma expansion, discharge neurologic severity measures, or 1-year mortality. Together, these findings suggest that in routine practice, SBP trajectories characterized by rapid early reduction and sustained lower levels (Cluster 2) are associated with increased renal risk without clear evidence of neurologic benefit.

Randomized trials established the biologic rationale for BP reduction after ICH—namely, the consistent relationship between elevated SBP and hematoma growth—and evaluated whether intensive lowering improves outcomes. INTERACT2 demonstrated that targeting SBP <140 mm Hg did not significantly reduce the primary composite endpoint of death or major disability, though ordinal functional outcomes suggested potential benefit^8^. ATACH-2 similarly found no improvement in the primary clinical endpoint with more intensive targets and raised concerns about adverse events with aggressive lowering^4,28^. Contemporary AHA/ASA guidance reflects these data and emphasizes careful implementation of acute BP lowering with attention to physiologic context and potential harms. Our findings complement this evidence base by showing that SBP trajectory “shape” and sustained exposure, not simply an initial target, may be clinically meaningful in practice. Whereas trials operationalize BP control through protocolized targets and time windows, clinicians often achieve BP goals through variable medication titration, differing monitoring fidelity, and evolving hemodynamics. The present analysis indicates that a persistently low SBP trajectory over days, rather than a transient early reduction alone, may identify a pattern of care associated with renal injury.

Several pathophysiologic mechanisms may explain the increased risk of AKI observed in patients with a rapidly dropped and stabilized SBP trajectory (Cluster 2). Many individuals with spontaneous ICH have long-standing hypertension, resulting in a rightward shift of renal autoregulatory thresholds. In this setting, rapid or sustained SBP reductions can lower renal perfusion pressure below the kidney’s autoregulatory range, predisposing to ischemic tubular injury—particularly in the context of critical illness, volume shifts, osmotherapy, and vasodilator use. In a secondary analysis of the ATACH-2 trial, patients experiencing large early SBP reductions (>90 mm Hg within the first 12 hours) had significantly higher rates of AKI, independent of baseline renal function and illness severity, suggesting that the *magnitude* of BP reduction itself contributes to renal injury^29^. Similarly, an observational cohort study of ICU patients with ICH demonstrated that aggressive SBP lowering was independently associated with AKI, even after adjustment for demographics, neurologic severity, and comorbidities, reinforcing the concept that renal hypoperfusion from intensive BP control represents a clinically meaningful tradeoff^30^. We also found that higher maximum nicardipine dose was independently associated with AKI. This aligns with secondary analyses from ATACH-2 showing that higher nicardipine exposure correlates with renal adverse events and AKI^29^. While nicardipine itself may not be directly nephrotoxic, higher doses likely reflect more difficult-to-control hypertension, greater BP variability, more severe physiologic derangements, or a need for stronger vasodilatory effect—each of which could contribute to renal vulnerability. Prior ICH studies often operationalize BP exposure using early maximal reductions (e.g., >90 mm Hg within 12 hours)^30^. Our trajectory-based approach instead captures the pattern of SBP over an extended period, which may better represent kidney risk as a cumulative hemodynamic exposure.

We did not observe statistically significant associations between SBP trajectory and hematoma expansion or downstream neurologic outcomes, a finding that likely reflects several non–mutually exclusive factors supported by prior literature. First, the temporal mismatch between exposure and benefit is critical: hematoma expansion occurs predominantly within the first few hours after symptom onset, and its likelihood declines substantially thereafter^31,32,33,34^. Consistent with this biology, randomized trials indicate that any potential benefit of BP lowering is driven by early SBP reduction, rather than prolonged maintenance of very low SBP; both INTERACT-2 and ATACH-2 achieved early BP separation but did not demonstrate improved clinical outcomes with sustained intensive control^8,28,35^. Thus, if both trajectory groups in our cohort achieved substantial SBP reduction within the first 12 hours, the incremental neurologic benefit of maintaining SBP <140 mm Hg over subsequent days would be expected to be limited. Second, measurement constraints likely attenuated detectable associations: hematoma expansion was assessed as a binary outcome based on radiology reports rather than volumetric quantification, an approach known to reduce sensitivity to modest yet clinically meaningful changes and bias results toward the null^36,37,38^. Finally, competing determinants of neurologic recovery likely outweighed the influence of SBP trajectory; baseline neurologic severity and age are consistently the strongest predictors of functional outcome and mortality after ICH, as reflected in established prognostic models incorporating admission NIHSS and ICH score, often eclipsing the effects of subsequent physiologic management once early stabilization has occurred (Hemphill et al., 2001; Cheung & Zou, 2003; Weimar et al., 2003)^17,39,40,^.

The observed protective association between 23.4% sodium chloride and AKI should be interpreted cautiously. Hypertonic saline is typically used in patients with elevated intracranial pressure or severe cerebral edema—indicating confounding by indication in either direction is plausible. Moreover, hyperosmolar therapy can influence urine output, serum creatinine dynamics, and fluid balance in complex ways that may not be fully captured by binary exposure coding. The parallel finding of higher mortality among those receiving 23.4% saline supports the notion that this variable primarily tags greater neurologic severity, rather than a causal protective renal effect.

Strengths of this study include the availability of high-resolution, longitudinal SBP data over the first 7 days of ICU admission, rigorous case verification through nurse-led manual chart review, exclusion of patients with advanced CKD to minimize baseline renal confounding, and application of a validated shape-based (*k*-Shape) clustering approach that captures clinically interpretable SBP trajectory phenotypes rather than relying on single time-point measurements. Several limitations merit consideration. The single-center, retrospective design introduces the potential for residual confounding and limits generalizability; however, we mitigated confounding by indication through multivariable adjustment for demographic factors, baseline neurologic severity, admission SBP, baseline renal function, antihypertensive exposure, maximum nicardipine dose, and prespecified nephrotoxic therapies, and by focusing on trajectory patterns rather than protocolized BP targets. BP measurements were derived from heterogeneous sources but to reduce measurement bias, we applied a standardized hourly sampling algorithm, avoided duplicate timestamps, and prioritized noninvasive measurements when simultaneous readings were available. Exclusion of short hospitalizations (LOS <5 days) may introduce survivorship bias; however, this criterion was intentionally selected to ensure adequate exposure assessment for SBP trajectories and AKI risk during the acute and subacute phases of care. AKI ascertainment may be influenced by documentation practices, fluid management, and catheter use; to address this, we employed standardized KDIGO criteria incorporating both serum creatinine and urine output and verified urine output data by documented indwelling catheter placement.

Prospective studies should evaluate whether trajectory-informed BP strategies can reduce AKI without increasing hematoma expansion, ideally integrating early hematoma biology and organ perfusion metric. Pragmatic trials or emulation studies could test conservative vs sustained-intensive approaches in subgroups at highest renal risk, and could incorporate medication dose-time integrals (e.g., nicardipine exposure) as mechanistic mediators.

## Data Availability

Please contact author with requests related to data included in study.

## Notes

### Competing Interest Statement

The authors have declared no competing interest.

### Funding Statement

This work has been produced with no external funding sources, all authors decline financial relationships to work.

### Author Declarations

This study was approved by the University of Rochester IRB Study #: STUDY00009896

